# Temporal and climatic drivers of uncomplicated malaria in Ghana: A Regional Generalised Additive Model analysis

**DOI:** 10.64898/2026.06.06.26355054

**Authors:** Edward Akurugu, Timothy Awine, Baba Seidu, Nana Yaw Peprah, Wahjib Mohammed, Paul Boateng, Paul Hilarius Asiwome Kosi Abiwu, Sheetal Prakash Silal

## Abstract

**Background:** Malaria remains a major public health challenge in Ghana, despite recent reductions in cases due to various interventions. The endemicity of the disease varies across regions, influenced by diverse seasonal and temporal factors that support mosquito proliferation and malaria cases. This study used a Generalised Additive Models to explore the impact of weather conditions on malaria cases in Ghana.

**Methods:** Generalised Additive Models were used to examine the nonlinear effects of weather conditions on malaria cases. Monthly aggregated malaria cases from the District Health Information Management System II and average monthly rainfall and temperature data from the Ghana Meteorological Agency were analysed, covering 2012 to 2023. Regional Generalised Additive Models incorporating weather variables were developed, fitted, and validated against observed data using model diagnostics to identify the most suitable model for each region.

**Results:** The analysis revealed complex temporal patterns in malaria cases across Ghana, influenced by seasonal and long-term trends. Regions constituting the Coastal and Transitional Forest zones exhibited bimodal peak malaria seasons, while the Guinea Savannah showed a unimodal peak. Significant interactions between rainfall and temperature were identified, particularly in the Eastern region, where higher rainfall combined with temperatures around 27–28 °C were associated with higher malaria cases, reflecting the complex and region-specific nature of meteorological influences.

**Conclusions:** The findings point to the dynamic and heterogeneous nature of malaria caseloads in Ghana, emphasising the need for region-specific control strategies tailored to local climatic conditions. A key recommendation is the systematic integration of meteorological data into the National Malaria Data Repository to enable continuous monitoring of climatic influences and support timely, evidence-based intervention decisions. Future research should incorporate socio-economic factors, intervention coverage data, vector surveillance, and demographic characteristics into mathematical modelling frameworks for a more comprehensive understanding of malaria cases in Ghana.

## Background

Malaria persists as a significant public health issue in sub-Saharan Africa (SSA), despite notable progress in reducing its burden. In 2024, an estimated 282 million cases were recorded across eighty endemic countries, with 610,000 deaths globally [1]. Two years earlier, the African region had borne a disproportionately high share of the global burden, accounting for 94% of malaria cases and 95% of malaria deaths. Specifically, children under five in Africa suffered 80% of these deaths [2]. Furthermore, malaria during pregnancy poses risks to the health of both the mother and baby, potentially leading to poor health outcomes, including death [3–5].

Effective malaria control strategies, such as indoor residual spraying (IRS) and insecticide-treated nets, among others [6–8], are known to have contributed significantly to the burden reduction. However, the 2023 WHO malaria report highlighted an increase of 5 million cases in 2022 compared to 2021, primarily in the WHO Africa region [2]. These trends have been consistent over the years in the WHO Africa region, highlighting the ongoing challenges in the fight against malaria, especially in the presence of spatial heterogeneity and temporal changes [2, 9, 10].

In Ghana, malaria is endemic nationwide, posing a constant risk to the entire population [11, 12]. Notwithstanding the successful control efforts, the country remains among the fifteen highest malaria-burden countries globally, accounting for approximately 2.5% of cases and 1.9% of deaths [13, 14]. As part of the WHO’s Global Technical Strategy for Malaria (2016–2030), Ghana’s National Malaria Elimination Program (NMEP)—formerly known as the National Malaria Control Program (NMCP)—aims to eliminate malaria by leveraging the National Malaria Elimination Strategic Plan (NMESP) from 2024 to 2028 [15]. However, the effectiveness of these strategies is influenced by the country’s diverse environmental conditions and seasonal variations [12, 14].

Malaria prevalence in Ghana varies throughout the year due to regional differences in climate and environmental conditions, with a significant increase during the rainy seasons [16]. This variability showcases the complex interplay between climate and malaria transmission in the country. Climate change and climate variability significantly affect the life cycle and transmission of malaria vectors by altering rainfall patterns and increasing the frequency of flooding and drought events [17]. Increased rainfall creates more breeding sites for mosquitoes, leading to a rise in malaria transmission, while intense rainfall may temporarily reduce mosquito populations by flushing early-stage larvae [18–24]. Higher temperatures enhance transmission by shortening the duration of parasite growth in mosquitoes and affecting their development, reproduction, survival, and biting rate [17, 25].

Climate variations play a key role in shaping regional malaria transmission patterns in Ghana [12, 26–30]. However, studies have reported conflicting relationship between climate and malaria incidence. This relationship appears to be nonlinear, characterised by spatio-temporal trends and seasonal variations across different regions [31, 32]. Similarly, studies from other countries have shown that the relationship between climate variables and malaria transmission is complex, with these factors, among others, impacting the disease in a nonlinear fashion [33, 34].

Several studies have examined the relationship between climate and malaria in Ghana, offering important but limited insights. Awine and colleagues analysed district-level malaria cases alongside rainfall and temperature using time-series and transfer function models, showing that climate effects were zone-specific and time-lagged [31]. However, their reliance on linear methods did not capture nonlinear dynamics. In a spatial analysis, Oppong and colleagues combined climatic and environmental covariates with georeferenced survey data to produce fine-scale malaria risk maps for Greater Accra. While they successfully identified high-risk areas, their focus was restricted to a single region and to prevalence surfaces rather than temporal climate–malaria relationships [35].

Together, these contributions highlight a clear gap: limited studies have employed flexible, nonlinear modelling frameworks capable of simultaneously capturing and quantifying region-specific climate effects on malaria transmission across Ghana at a large scale.

Building on this foundation, this study uses a Generalised Additive Model (GAM) to investigate the nonlinear effects of climate covariates on malaria across all ten former regions of Ghana [36, 37]. This approach enables a deeper understanding of the diverse dynamics and interactions between meteorological variables and malaria at the regional level. Hence, focusing on temporal heterogeneity of malaria, this study provides novel insights that can guide the development of targeted intervention strategies, accelerating efforts towards malaria elimination across the country. To our knowledge, this is the first study to apply GAM across all the ten former regions of Ghana, highlighting its potential to significantly inform policymaking and intervention planning in each of the different regional settings.

## Materials and methods

### Study setting and population

Ghana, located in the West African sub-region, borders Burkina Faso to the North, the Atlantic Ocean and Gulf of Guinea to the South, Togo to the East and Côte d’Ivoire to the West [11]. The 2021 Population and Housing Census (PHC) estimates indicate Ghana’s population to be 30.8 million people [38], with a land mass of approximately 238,533 *km*^2^ [39], which is currently divided into sixteen administrative regions as indicated in **Fig 1** [11, 38]. These administrative regions are also categorised into three ecological zones (Coastal, Transitional Forest, and Guinea Savannah) due to differences in rainfall, temperature, and vegetational patterns [31, 40, 41].

**Fig 1.**
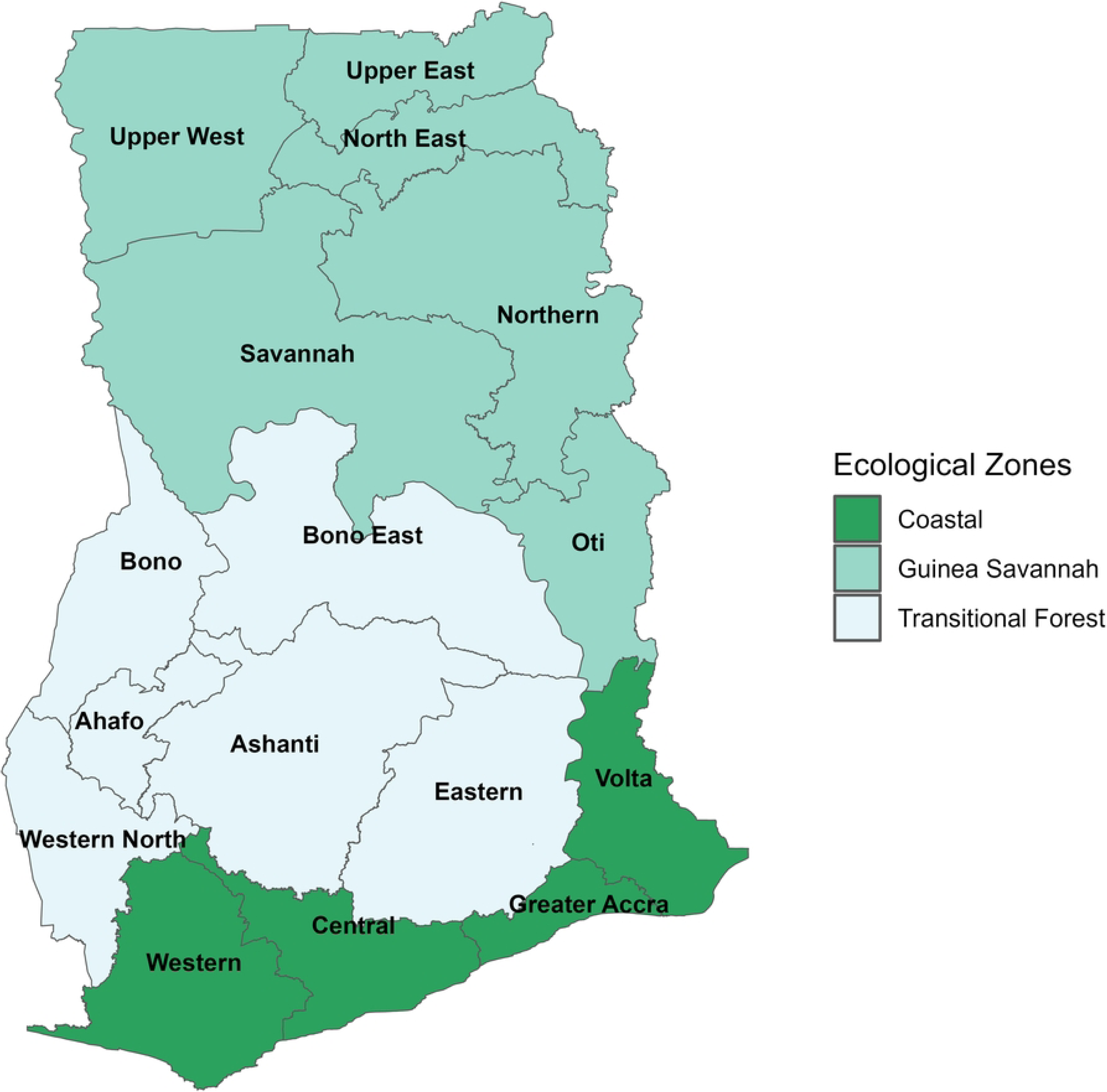
Map of Ghana showing administrative regions across ecological zones. (Data source: https://gadm.org/download_country.html)

Under the current sixteen-region administrative structure, the Coastal zone comprises four regions, while the Guinea Savannah and Transitional Forest zones each have six regions [40]. However, this study analyses the former ten region of Ghana, which are grouped as follows: the Coastal zone comprises three regions (Central, Greater Accra, and Western), the Transitional Forest zone comprises four regions (Brong Ahafo, Ashanti, Volta, and Eastern), and the Guinea Savannah zone comprises three regions (Upper East, Upper West, and Northern). The Coastal zone is characterised by a sandy coastline with many streams and rivers. The Transitional Forest zone is dominated by heavily forested areas with streams and rivers, while the Guinea Savannah zone is characterised by grasslands and scattered trees (savannah vegetation), traversed by the White and Black Volta rivers [40].

### Study design and period

This study seeks to analyse the nonlinear relationship between malaria cases and meteorological factors using historical data sets. Malaria case data and meteorological data covering January 2012 to December 2023 were compiled and analysed to explore temporal and climatic patterns.

## Data description

### Malaria case data

In Ghana, a common platform known as the District Health Information Management System II (DHIMS2) is used at all levels of healthcare to capture malaria (morbidity and mortality) data from all the health facilities (public and private) at the various levels, including the community, sub districts, and districts. Malaria case data captured on the DHIMS2 platform are aggregated daily, weekly, monthly, quarterly, and annually at these levels (community, sub districts, and districts) and then compiled at the regional level before reaching the national level.

The creation of six new regions in 2019 by a legislative instrument posed a problem regarding obtaining the relevant historical dataset from 2012–2023 for these new regions. To circumvent this, 2019–2023 datasets for the six new regions (Western North, Ahafo, Bono East, Oti, Savannah, and North East; **Fig 1**) were combined with datasets from the regions from which they were carved. For example, the datasets for Western North and Oti regions were merged with the data from the Western and Volta regions, respectively. Similarly, the datasets for the Ahafo and Bono East regions were merged with data from the previous Brong Ahafo region, while the datasets for Savannah and North East regions were combined with data from the previous Northern region.

In this study, we used the monthly regional malaria case surveillance data from January 2012 to December 2023, obtained through the DHIMS2 platform and consolidated as described above. These data include all patients (including children, adults, and pregnant women) that visited health facilities, and presenting with malaria symptoms, were diagnosed with malaria either through a rapid diagnostic test (RDT) or microscopy.

### Meteorological data

The study used monthly regional meteorological data from January 2012 to December 2023. The historical meteorological data used in the study include rainfall and average temperatures (minimum and maximum). These datasets were obtained from the Ghana Meteorological Agency (GMeT). Similar to the malaria case data, to address the limitations of acquiring monthly datasets from 2012–2023, we consolidated the meteorological datasets for the six new regions created in 2019 (Western North, Ahafo, Bono East, Oti, Savannah, and Northeast) with the data from their parent regions. It is recognised that the consolidation approach may inevitably introduce some spatial approximation, particularly in regions with distinct climatic conditions relative to their parent regions, and this constitutes a limitation of the study.

### Inclusion and exclusion criteria

The study encompassed all categories of persons (children, adults, and pregnant women) who visited public or private regional health facilities. Patients were included if they were assessed and confirmed with uncomplicated malaria through RDTs or microscopy, and had their data recorded in the DHIMS2 platform during the study period (2012–2023). Uncomplicated malaria was chosen for the study because it represents the majority of cases, provides a more accurate picture of the overall malaria caseloads, and allows for consistent comparison across regions and time periods due to standardised treatment protocols. Hence, examining uncomplicated malaria, we seek to identify patterns that can inform preventive strategies and early intervention programs, potentially reducing progression to severe malaria cases.

Severe or complicated malaria cases were excluded, as were suspected malaria cases without laboratory confirmation and patients whose data were not captured on the DHIMS2 platform. This approach ensures that the study focuses on a more uniformly diagnosed and documented form of malaria. Uncomplicated malaria is generally more consistently diagnosed due to standardised treatment protocols and less variability in clinical judgement compared to severe or complicated cases [42–44]. Severe malaria cases often involve more complex diagnostic and treatment decisions, which can vary significantly between clinicians and facilities. This variability can introduce additional uncertainty into the analysis, making uncomplicated malaria a more reliable focus for studying seasonal and regional trends.

### Data management, quality control and analysis

Under this study, the malaria and climate datasets were compiled and structured in Microsoft Excel prior to importation into R for analysis. The meteorological dataset was accessed from GMet for research purposes on 15th May 2024, while with the malaria surveillance data, it was accessed from the DHIMS2 platform through the NMEP on 27th May 2025. All apparent missing entries were cross-checked with the respective data providers, and no missing data were identified at the point of analysis. Monthly case counts and meteorological variables were inspected for implausible values and reporting inconsistencies across all regions and time points, and no such irregularities were detected. All regional datasets were obtained and used exclusively in aggregated anonymised form. Hence, no individually identifiable participant information — including names, personal identifiers, or facility-level records — was accessed, stored, or used during or after data collection. It is acknowledged that variations in testing rates and facility reporting completeness across regions and over time were not adjusted for, and this represents a limitation of the study.

Descriptive statistics including minimum, mean, maximum, standard deviation, and coefficient of variation were computed for malaria cases and meteorological variables at the regional level. Seasonality was assessed using the Kruskal-Wallis and Friedman tests, and the association between malaria cases and meteorological variables was modelled using GAMs as described below.

### Generalised additive model

GAM is an extension of linear regression and generalised linear models, with an additional layer in which the GAM models the nonlinear relationships among predictor variables and the response variable. In this study, we examined the impact of meteorological variables on malaria case counts (uncomplicated malaria) using a GAM by considering long-term trend, seasonality, and temporal variations. Consider *Y*_*it*_ to be the monthly malaria case counts and *E*_*it*_ as the expected counts in region *i* and month *t*, where *i* = 1, 2, ⋯, *I* and *t* = 1, 2, ⋯, *T*. Also, since the dependent variable (uncomplicated malaria cases) is a count variable, it is important to consider count regression models in the modelling framework.

The baseline model for modelling count variables, such as malaria cases, is the Poisson regression model. However, during the modelling process, datasets that are counts present lots of challenges, especially when the data are characterised by a preponderance of excess zeros or are overdispersed [45]. Moreover, the assumptions of equidispersion and independence make it inappropriate to handle count data with the above-mentioned characteristics due to dependencies in individual responses, heterogeneity, clustering, contagion, and non-inclusion of some causal factors associated with the disease [46, 47].

The problem of overdispersion when modelling count data can be addressed by considering fitting the data using different families of the Poisson regression models (such as the quasi-Poisson, negative binomial, among others). The association between malaria transmission and climate and environmental factors is quite debatable [48, 49], and the impacts of these environmental variables are nonlinear and vary across their ranges [34, 50]. GAMs have been increasingly applied in epidemiological studies to capture such nonlinear effects of climate on vector-borne diseases. For example, [51] used GAM to assess weather sensitivity of under-five malaria in Ghana, and [52] applied GAM to model climatic influences on malaria transmission in northeastern India. These applications demonstrate the flexibility of GAMs in modelling complex temporal and environmental relationships, providing a strong methodological foundation for their use in this study.

Accordingly, we used GAM to understand the intricate relationships between malaria and meteorological factors, fitting different families of count distributions (Poisson, quasi-Poisson, and negative binomial) to the full GAMs (S1 Table 1B in S1 Text) in each region. The full GAMs in each region served as benchmarks to determine the appropriate conditional distribution for fitting the observed data, based on the outcome of model selection criteria. Based on these criteria, the regional data from the full GAMs were found to follow a quasi-Poisson distribution, which was subsequently used for fitting the regional data. The general GAM is given as follows:

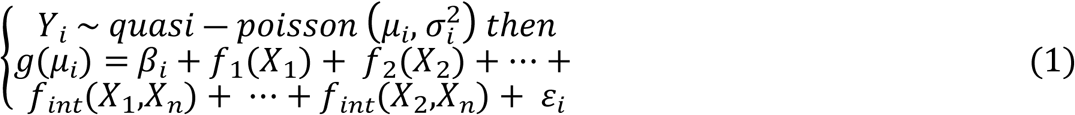

where from Eq (1), *y* is the response variable (uncomplicated malaria cases), *g* is the log–link function that relates the mean *μ* to the predictors, and *ε*_*i*_ is the residual error term for observation *i*. Also, the parametric term *β*_*i*_ in Eq (1) is the intercept and *X*_1_, *X*_2_,⋯,*X*_*n*_ are the explanatory variables.

Moreover, the non-parametric terms in Eq (1), namely *f*_1_, *f*_2_,⋯,*f*_*n*_ and *f*_*int*_ capture the smooth main effects and smooth interaction effect, respectively. The full GAM model and the corresponding reduced models considered in this study are captured extensively (S1 Table 1A in S1 Text).

The regional data preparation was conducted in R version 4.5.2 before the onset of the formal analysis for the regional GAMs. Within the GAM framework, the **mgcv** package in R [53] was used to fit each regional GAM to the observed data.

### Model building strategy

The backward elimination strategy, as shown in Table 1 and **S1 Fig**, was used to build the GAMs (S1 Table 1A in S1 Text) considered in this study. The strategy involved first fitting the full GAM (S1 Table 1A in S1 Text) with all the non-parametric terms, correctly specifying the smoothing functions and regression splines. Subsequently, the full GAM was evaluated to determine whether some modifications regarding the adjustment of the basis dimension (*k*) were plausible. This process continues iteratively until the final parsimonious model that best fits the underlying data was achieved. The algorithm and conceptual workflow illustrated in Table 1 and **S1 Fig**, respectively, shows the steps involved in building the GAMs considered in this study.

**Table 1.**
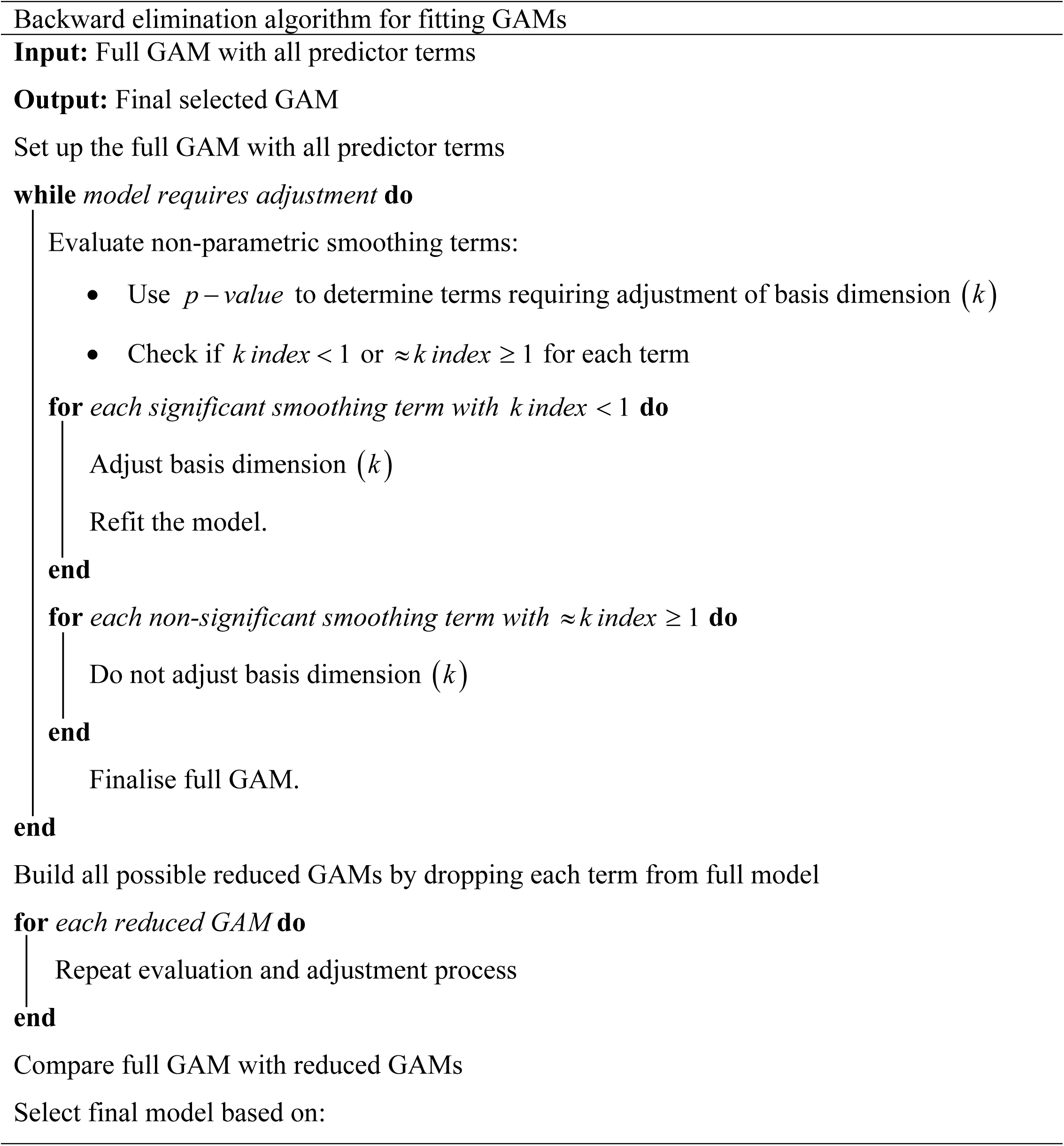
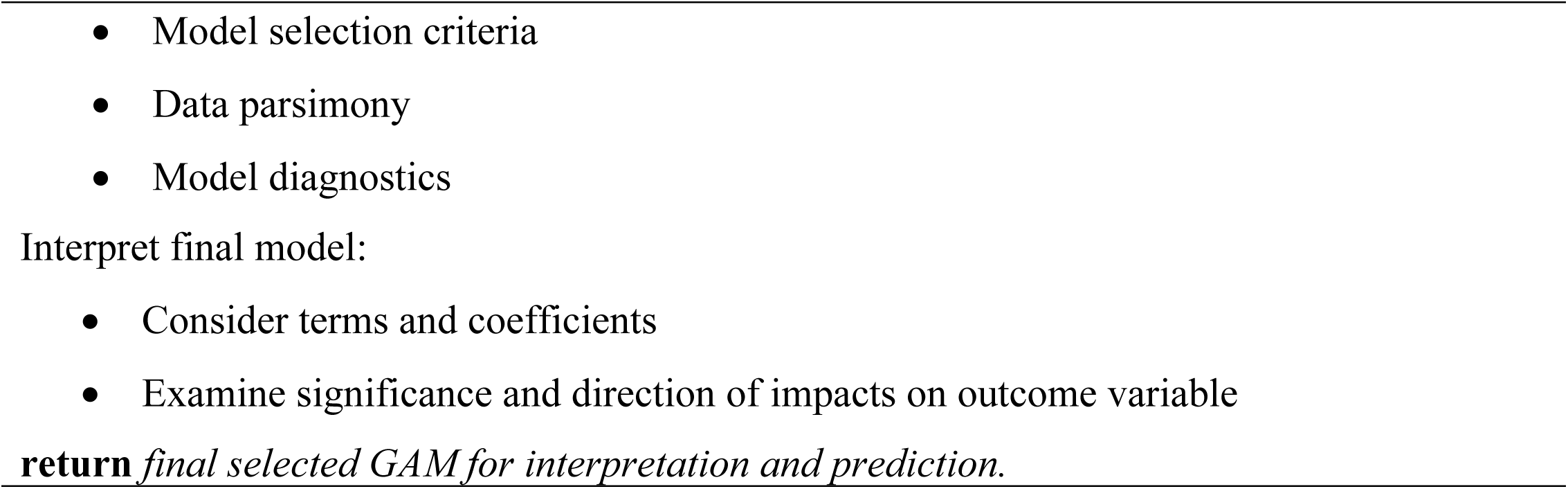
Model building process for all regional GAMs.

### Model diagnostics

The decision to settle on a model was made after it had been subjected to diagnostic assessment to evaluate its performance in real-life situations. In this study, several model diagnostic tools within the GAM framework, such as normal Q–Q plots and deviance residuals, were used to assess model performance. Also, model selection criteria (adjusted R–squared and explained deviance) were applied to determine whether the residual error terms associated with each regional model violated underlying assumptions, such as normality and randomness. The diagnostic results for the selected GAMs are presented in S5 Fig 1A–1C, providing visual confirmation of model adequacy and robustness.

## Results

### Summary analysis of malaria cases and meteorological variables

Table 2 presents the descriptive statistics of malaria cases and meteorological variables across the former ten regions of Ghana from 2012 to 2023. The table provides minimum, mean, and maximum values for malaria cases, average temperature, and rainfall in each region.

**Table 2.** Descriptive statistics of malaria cases and meteorological variables (2012–2023)

| Region | Uncomplicated malaria (n) |  |  | Average Temperature (°C) |  |  | Rainfall (mm) |  |  |
| --- | --- | --- | --- | --- | --- | --- | --- | --- | --- |
|  | Min* | Mean | Max* | Min* | Mean | Max* | Min* | Mean | Max* |
| Upper East | 10101 | 35210 | 95861 | 26.6 | 29.4 | 34.4 | 0 | 86.8 | 352.2 |
| Upper West | 4850 | 19235 | 50527 | 24.4 | 28.5 | 32.8 | 0 | 88.6 | 382.3 |
| Northern | 3398 | 28592 | 67538 | 23.4 | 29.4 | 34.2 | 0 | 86.7 | 333.6 |
| Brong Ahafo | 20011 | 65710 | 125739 | 24.6 | 27.0 | 30.4 | 0 | 97.1 | 320.7 |
| Ashanti | 22020 | 52947 | 93468 | 24.6 | 27.5 | 30.7 | 0 | 125.0 | 398.3 |
| Eastern | 21129 | 50469 | 93167 | 25.3 | 27.8 | 30.4 | 0 | 108.3 | 296.0 |
| Volta | 8150 | 29906 | 61693 | 25.4 | 28.1 | 31.0 | 0 | 108.4 | 379.2 |
| Greater Accra | 7391 | 17590 | 26798 | 24.6 | 28.2 | 30.5 | 0 | 68.7 | 380.3 |
| Central | 14803 | 39160 | 66695 | 24.21 | 26.4 | 28.7 | 0 | 83.7 | 449.4 |
| Western | 15443 | 52487 | 94534 | 24.5 | 27.31 | 29.3 | 0 | 102.98 | 694.1 |

\*Min and Max: minimum and maximum values for the period, 2012–2023.

The observed data reveal substantial variations in malaria cases across the regions in Ghana. Brong Ahafo reported the highest mean number of malaria cases (65,710) and the highest maximum (125,739) cases over the study period. This finding aligns with similar studies such as [54], who identified the Brong Ahafo region as a high malaria prone area. In contrast, Greater Accra showed the lowest mean (17,590) and maximum (26,798) number of cases. Other regions, such as Ashanti, Western, and Eastern also reported high mean malaria cases (52,947, 52,487, and 50,469 respectively), indicating a substantial high malaria caseload.

Average temperatures across regions typically ranged between 26–29 °C. The Upper East, Northern, and Upper West regions experienced the highest maximum temperatures (34.4 °C, 34.2 °C and 32.8 °C, respectively), while the Central region recorded the lowest mean temperature (26.4 °C). Rainfall patterns varied, with all regions experiencing periods of no rainfall (minimum 0 mm). The Western region recorded the highest maximum rainfall (694.1 mm), while Greater Accra had the lowest mean rainfall (68.7 mm).

### Patterns of malaria and meteorological variables

The results from Table 2 show substantial differences between minimum and maximum values for all variables, suggesting seasonal or annual variations in both malaria cases and meteorological conditions. These variations are not clearly visible in the time series plots in Fig 2, although there are consistent yearly variations with regular peaks and troughs observed in each region over the study period for rainfall and average temperature.

**Fig 2.**
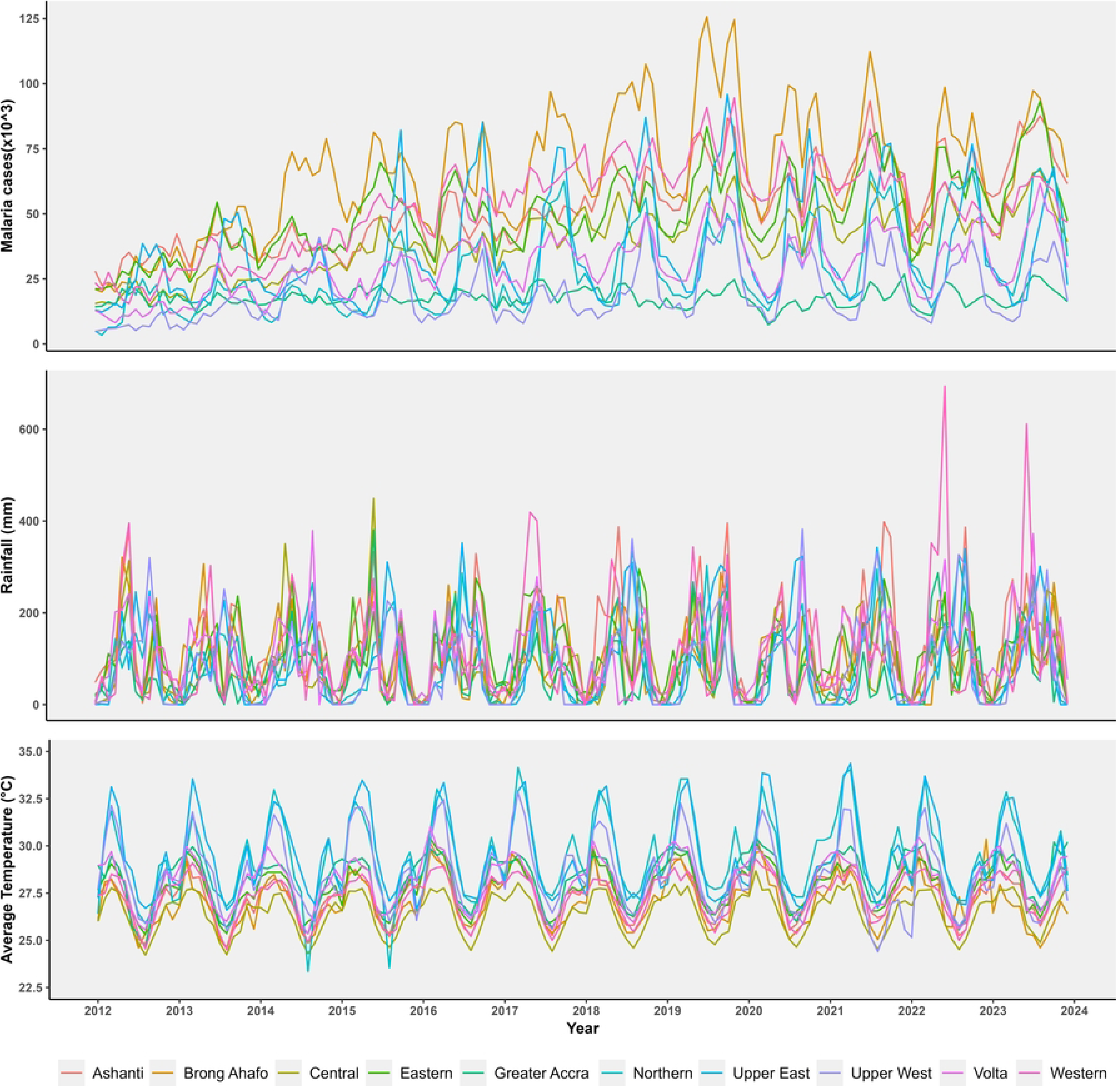
Patterns of regional malaria cases (top panel), rainfall (middle panel), and average temperature (lower panel) by year.

The malaria cases, as shown in the top panel of Fig 2, exhibits trends that are not easily discernible, with regular peaks and troughs for each of the regions. These trends can be best observed from the time series decomposition using locally estimated scatterplot smoothing (LOESS) in S3 Fig 1A. The seasonal malaria case series decomposition in S3 Fig 1B coincide with the patterns in **S2 Fig 1A-1J**, displaying seasonality. Regions in the Transitional Forest and Coastal ecological zones (Brong Ahafo, Ashanti, Eastern, Volta, Central, Greater Accra, and Western) exhibit two peak seasons of malaria compared to a single peak season observed in the regions constituting the Guinea Savannah zone (Upper East, Upper West, and Northern).

The monthly aggregated regional average temperature observations from Fig 3 range from 23–34 °C, which supports the malaria transmission range [32]. High temperatures typically occur between February and May while low temperatures are observed between June and October across all ten regions of Ghana.

**Fig 3.**
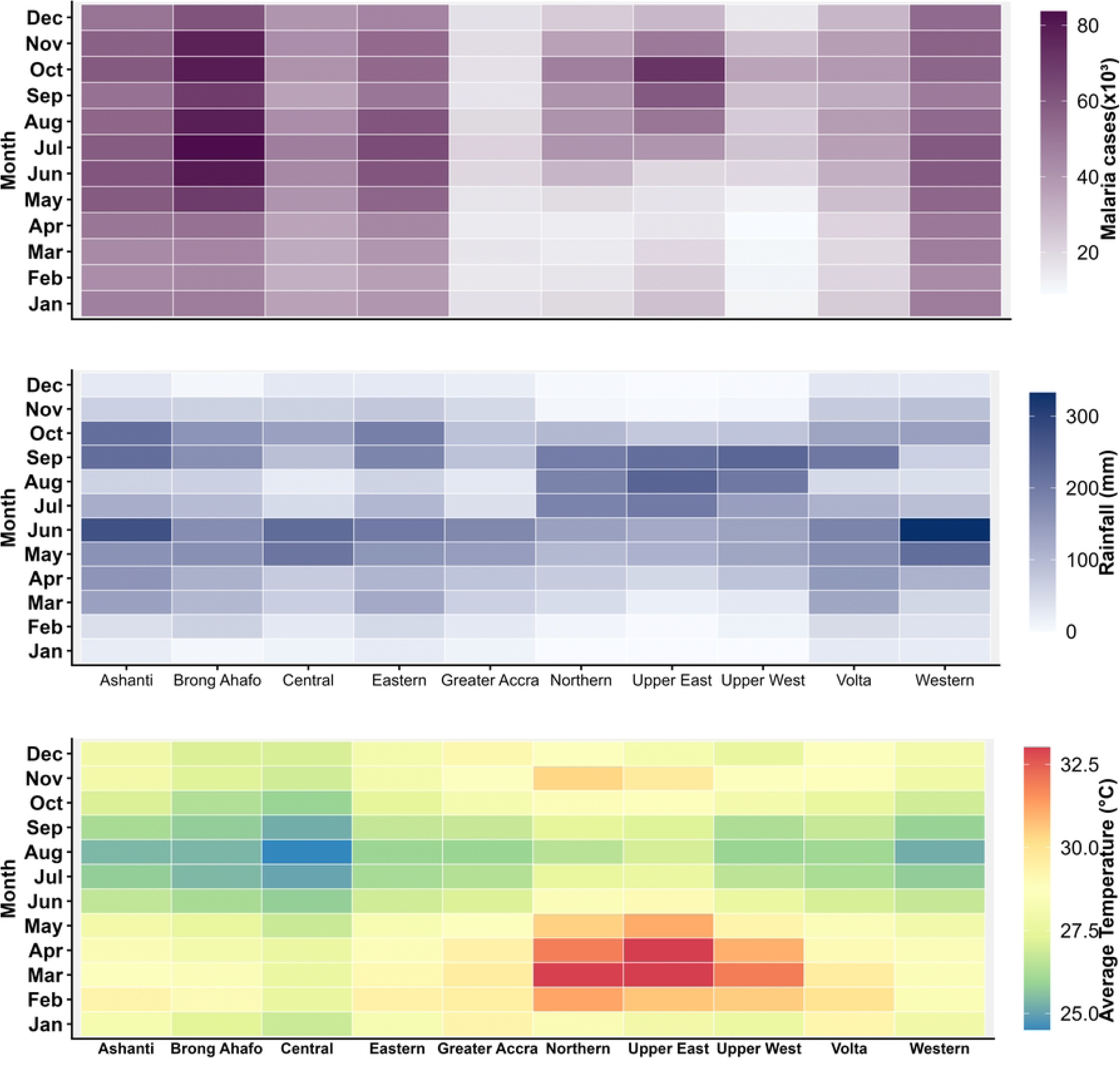
Patterns of regional monthly aggregated malaria cases (top panel), rainfall (middle panel), and average temperature (lower panel).

The patterns of rainfall vary across all regions of Ghana, typically ranging from 0 to 694 mm per month (Fig 2, S2 Fig 1A–1J, and Fig 3). The Guinea Savannah zone (Upper East, Upper West, and Northern) generally has a unimodal rainfall pattern, which falls in line with studies by NMEP and United States President’s Malaria Initiative [12, 55], with peak seasons occurring in August (Upper East) or September (Upper West and Northern) (S2 Fig 1A–1C). The Transitional Forest (Brong Ahafo, Ashanti, Volta, and Eastern regions) and Coastal zones (Central, Greater Accra, Western regions) tend to have a bimodal rainfall pattern (S2 Fig 1D–1F), which supports studies by NMEP and United States President’s Malaria Initiative [12, 55]. The first major rainfall peak occurs in June, with the second peak varying among regions: September for Brong Ahafo, Ashanti, and Volta; October for Eastern, Central, and Western, and September for Greater Accra.

The results in S3 Fig 1A show that the regional malaria cases in Ghana depict nonlinear trends. This indicates that the observed trends in malaria cases in each region are complex, requiring advanced models such as GAMs to capture these underlying patterns and other influencing factors, such as meteorological variables, that characterise the malaria data.

### Temporal trends and meteorological effects on malaria cases

The GAM was used to examine the temporal patterns of malaria across the various regions of Ghana, incorporating the influence of long-term trend (time), seasonality, average temperature, and rainfall. Eight models were fitted in each region with the best models in each of these regions selected using generalised cross validation, adjusted *R*^2^, and deviance explained. The results from the selected models observed in Table 3 reveal complex and region-specific patterns in the dynamics of malaria cases. These variations arise from the observed varying degrees of regional specific temporal complexities, interaction effects, seasonal dynamics, and differential impacts of meteorological factors observed in the estimated degree of freedom (*edf*) in Table 3.

**Table 3.**
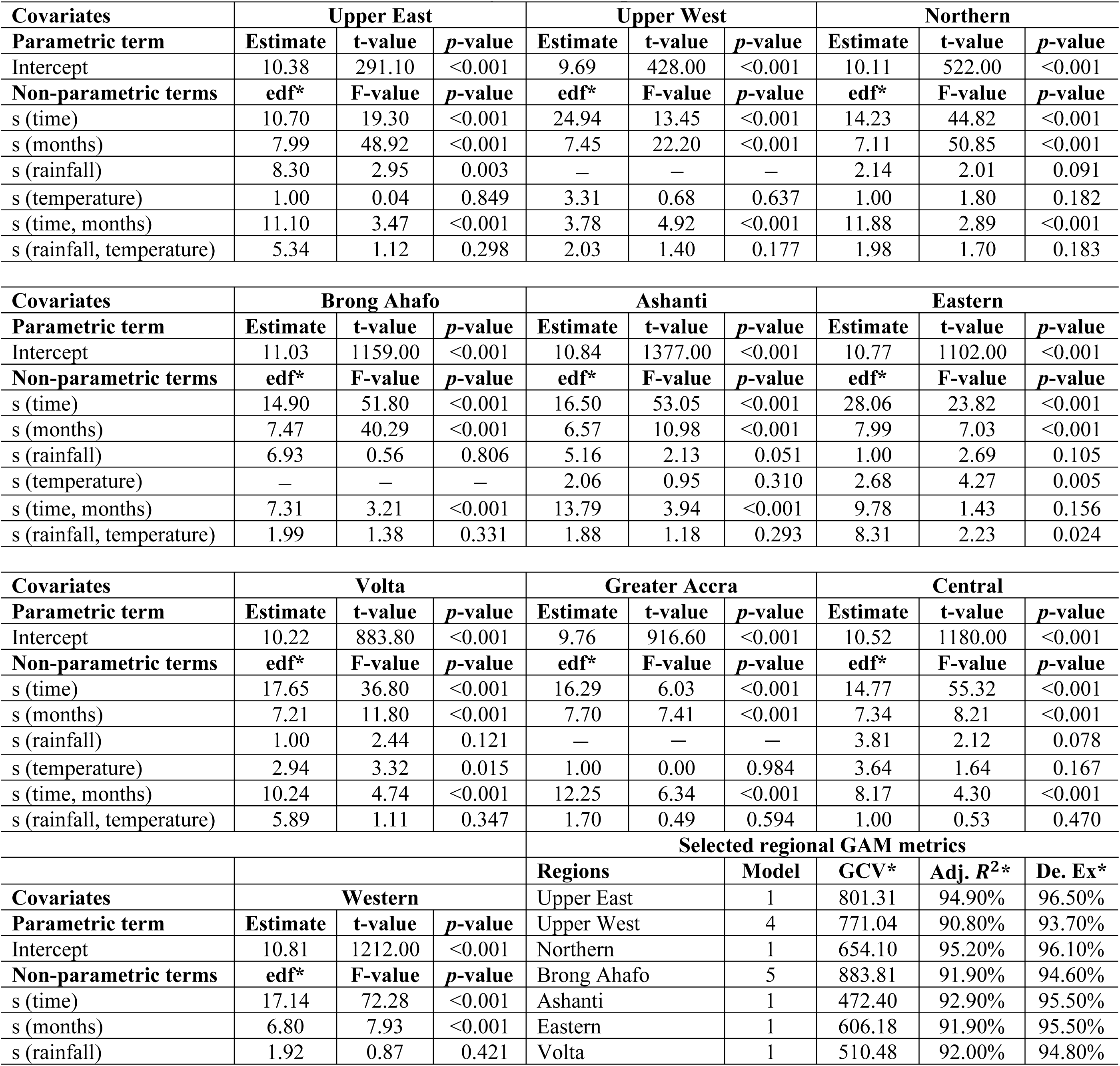
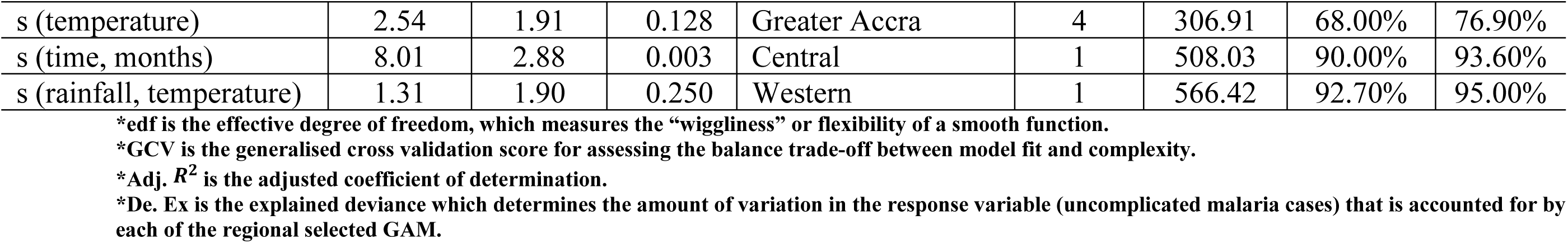
Estimated effects of meteorological and temporal factors on malaria cases in Ghana.

The model intercepts in Table 3 depict the log-transformed expected monthly malaria caseloads considering all other variables in the model are at their average values. The results of the intercepts (*p* = 0.001) show that all regions in Ghana tend to have a substantial level of malaria, culminating in the disease being considered a major public health concern in most studies [12, 55]. This finding aligns with numerous studies that have stressed the significant impact of malaria in the country [56, 57].

Besides the significance of the model intercepts, the analysis indicates significant regional variations in the influence of meteorological factors on malaria in the country. Long-term trends and seasonality were identified as contributors to the prediction of malaria cases across all the regions. Despite this, apart from the Eastern region, long-term trend and its interaction with seasonality were identified as key predictors of malaria cases (Table 3), confirming the strong temporal and seasonal patterns of malaria cases in these regions.

Results in Table 3 also point to the varying levels of significance for meteorological variables, highlighting the importance of environmental conditions in malaria cases. Rainfall significantly affects malaria cases in the Upper East region (*p* = 0.003); however, the effects in Ashanti (*p* = 0.051), Central (*p* = 0.078) and Northern regions (*p* = 0.091) are marginally significant at 10% level.

Average temperature substantially affects malaria cases in the Eastern region (*p* = 0.005) and Volta region (*p* = 0.015). Regarding the interaction between the rainfall and average temperature, the effect was found to significantly influence malaria case patterns in the Eastern region (*p* = 0.024).

Supporting these findings, the smoothing terms in Fig 4 and S4 Fig 1A–1G indicate that the effects of all the regional long-term trends and seasonality tend to increase as time progresses, with some level of certainty in the estimates. However, the variability (wiggliness) in the trend was much more pronounced in the Upper West and Eastern regions, revealing the complexities associated with capturing these regional trends. Reported malaria cases generally decrease from January until around April and towards the last two months of the year across the various regions (Fig 4 and S4 Fig 1A–1G).

**Fig 4.**
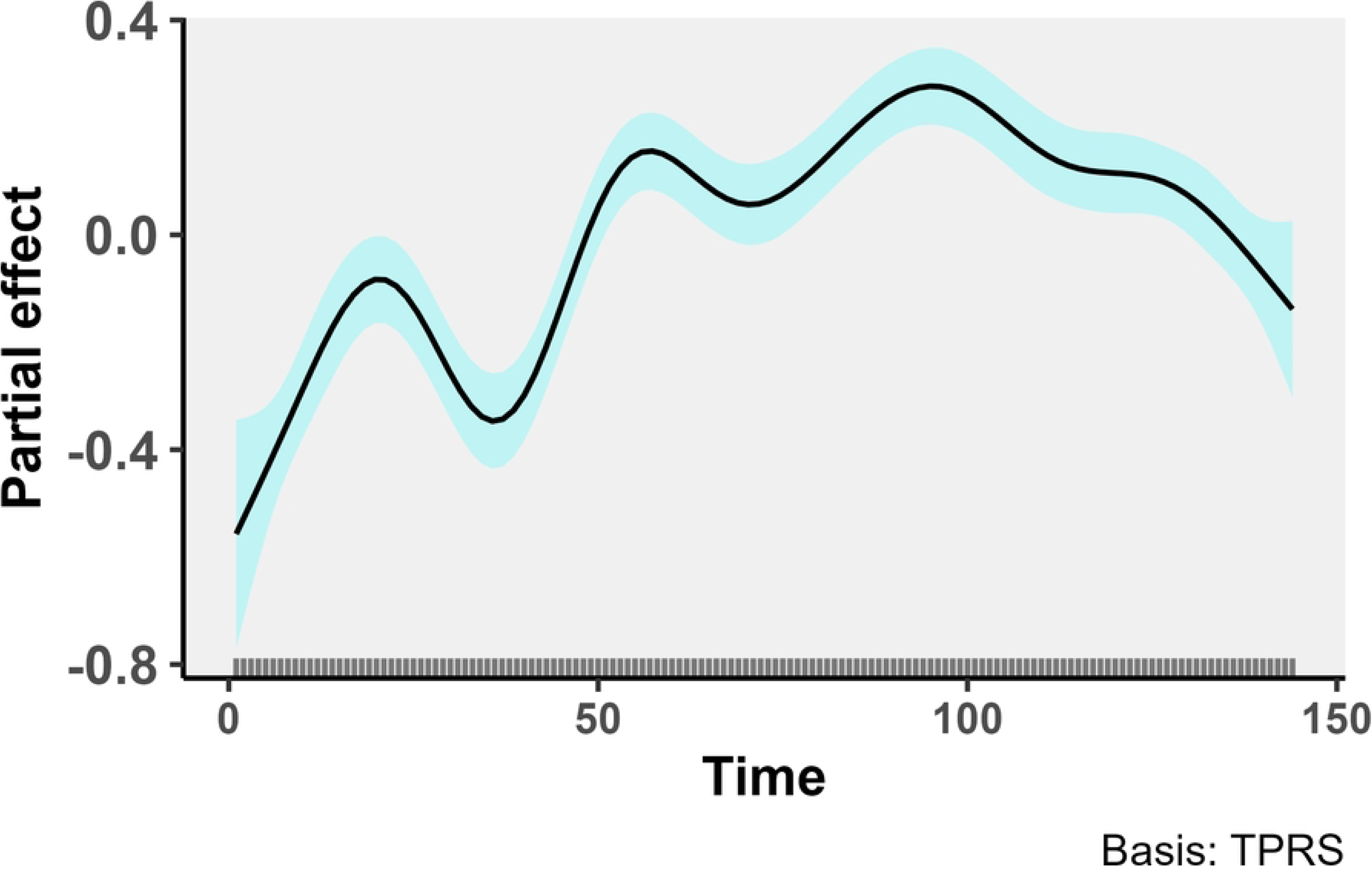
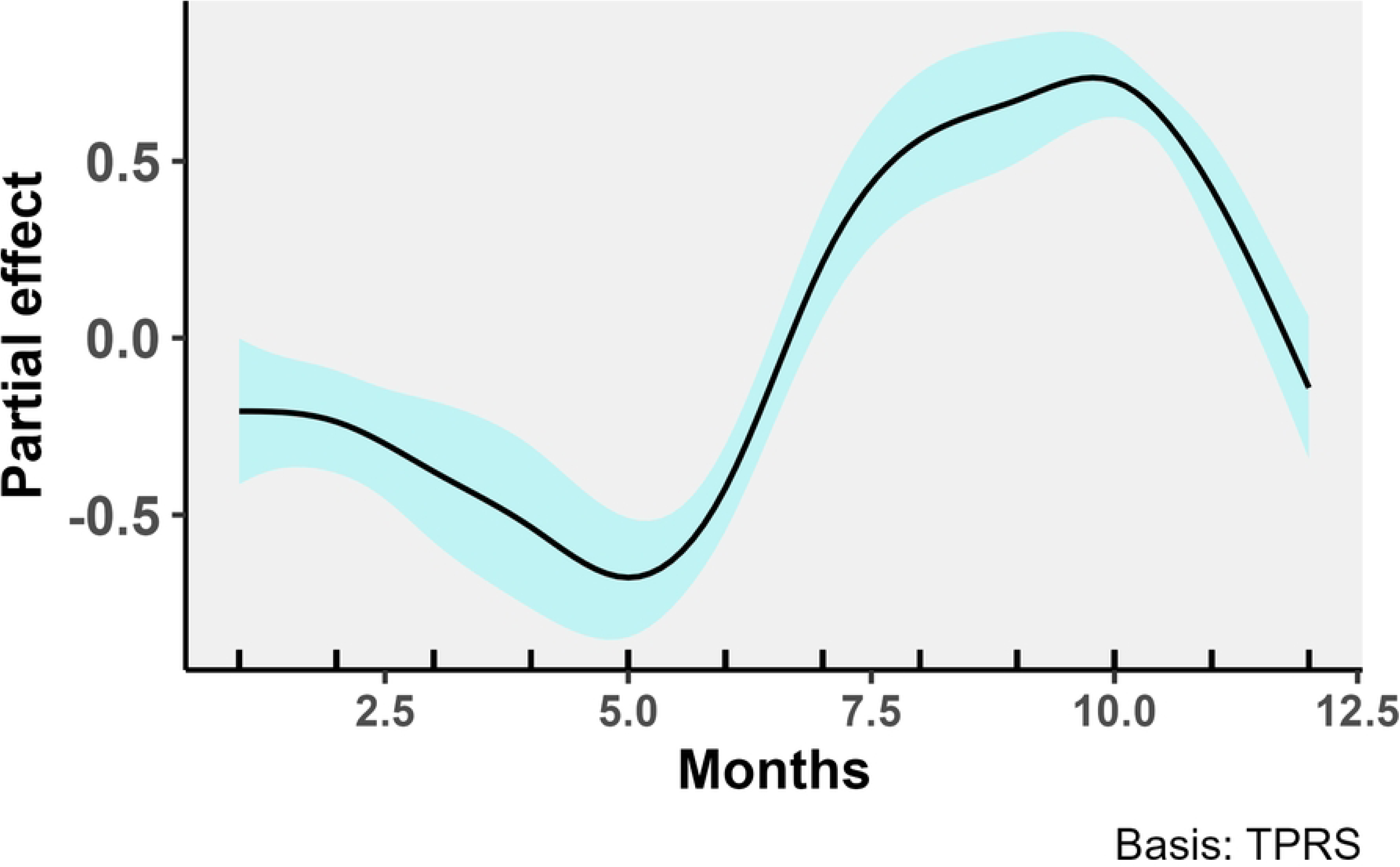
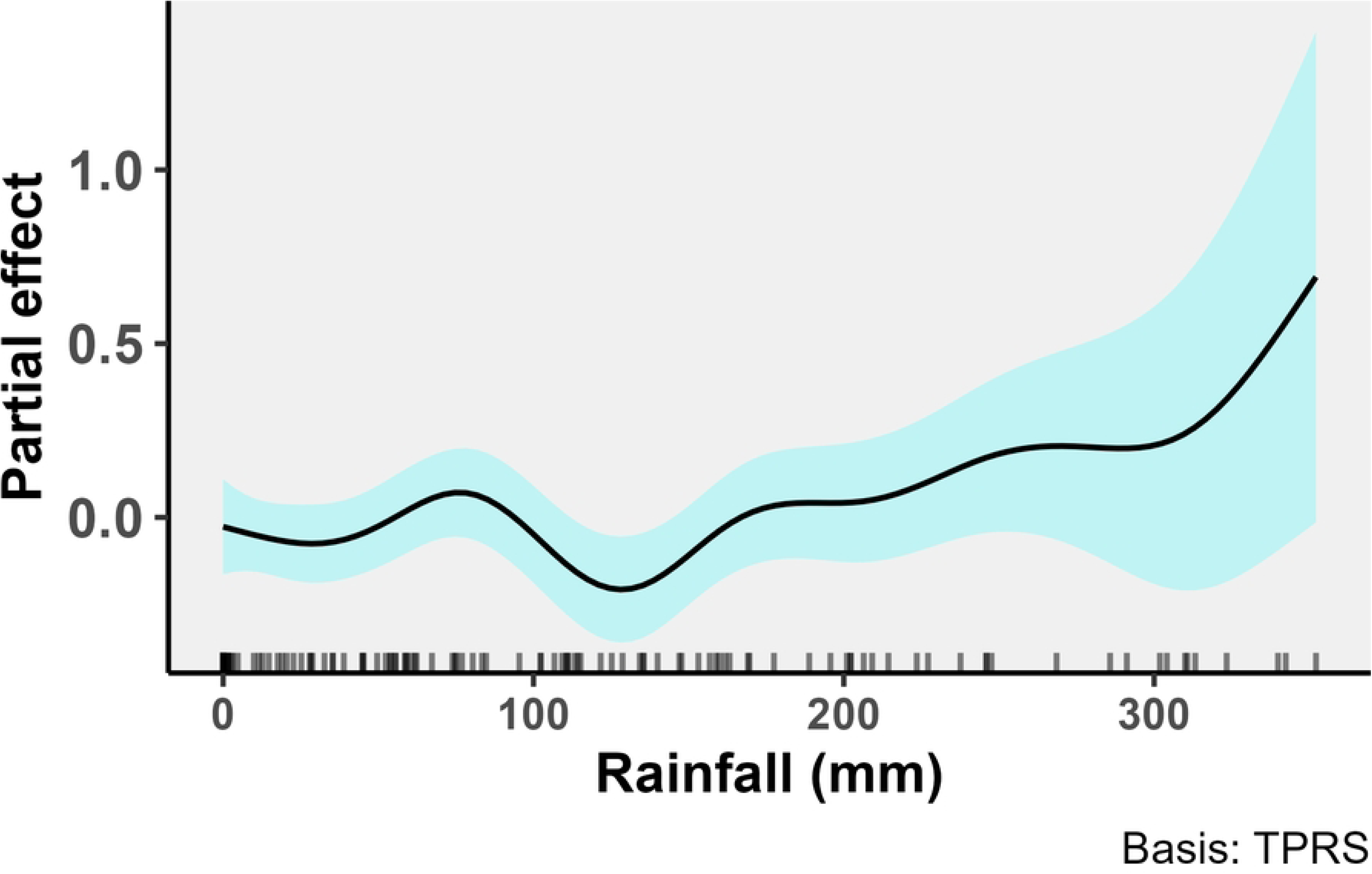
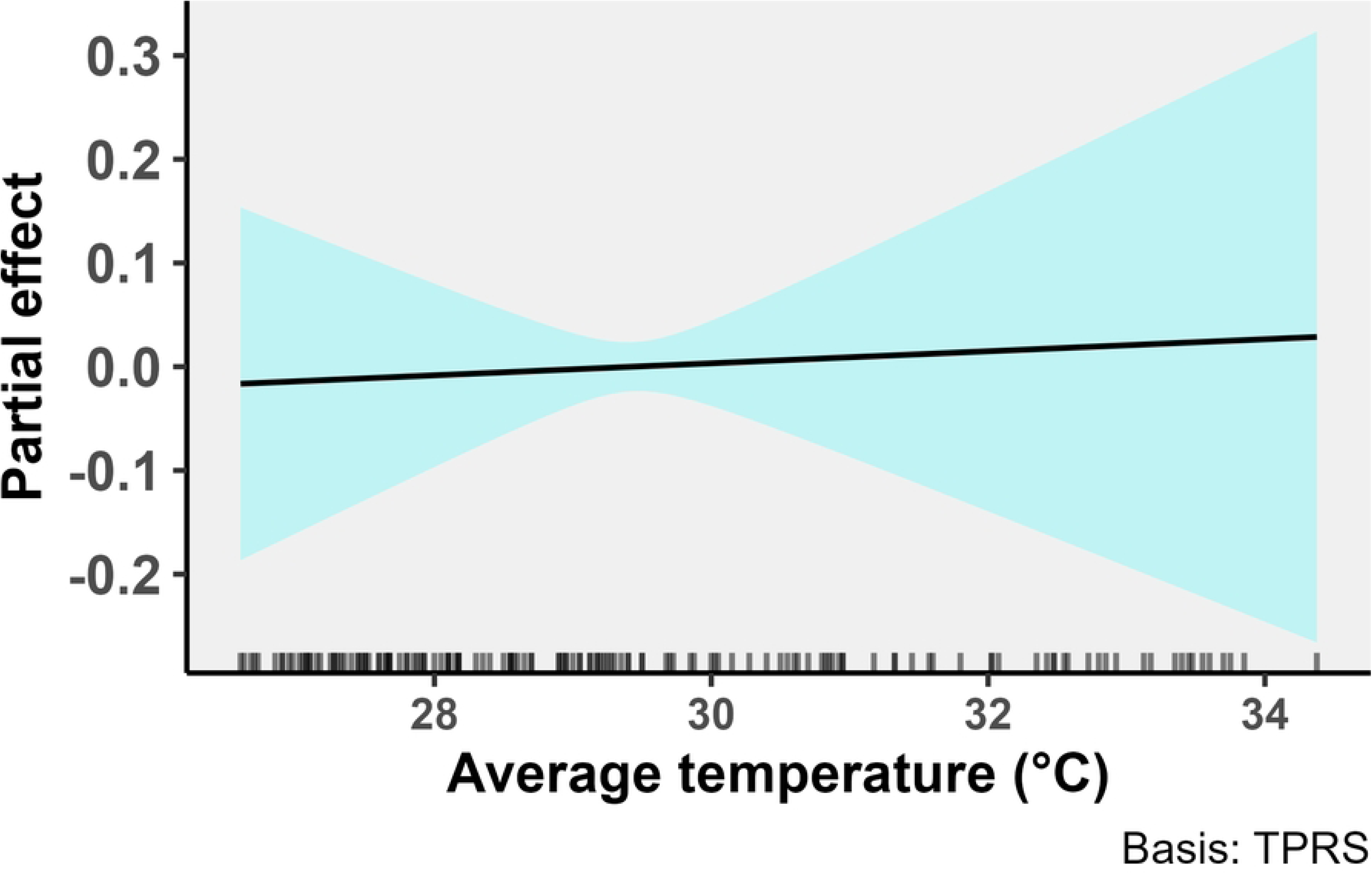
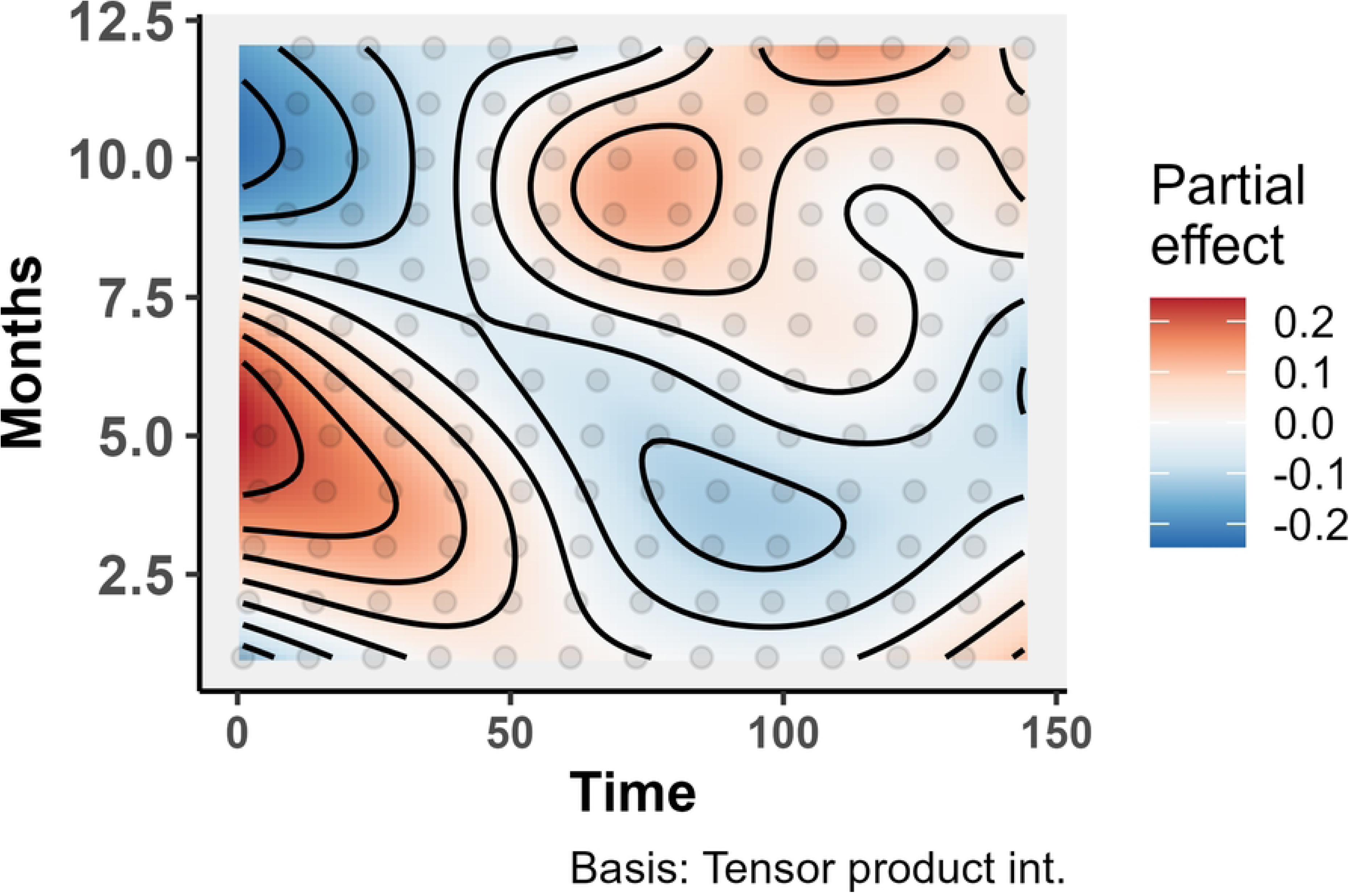
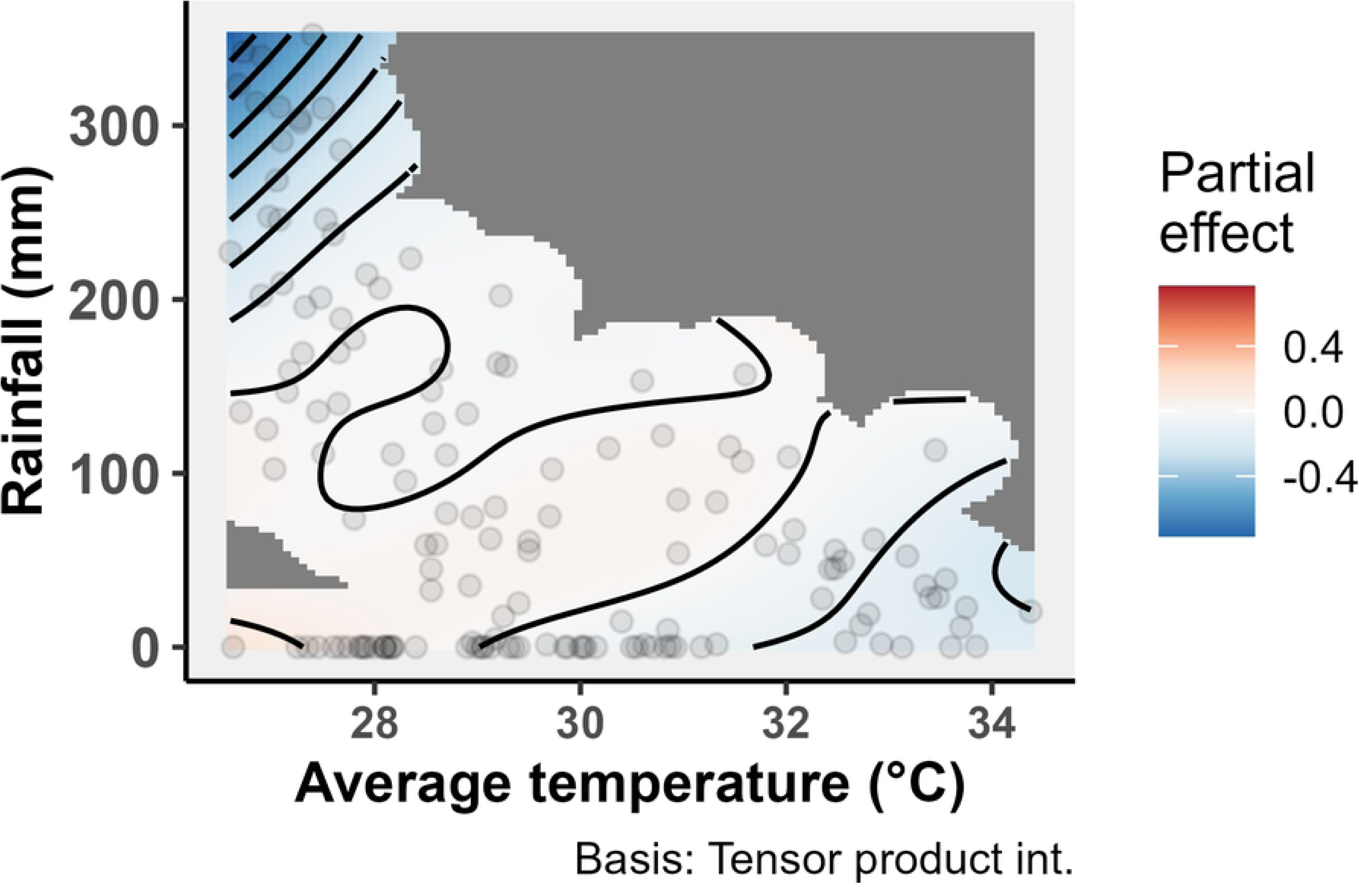
Partial effects of covariates (long-term trend [time], seasonality [months], rainfall, average temperature, interactions between long-term trend [time] and seasonality [months], and interactions between rainfall and average temperature) on malaria cases in the Upper East region. The cyan colour around the black lines in the plots represents the estimated uncertainties from the 95% confidence interval for the period 2012–2023.

The nonlinear relationship between rainfall and malaria cases varies across the regions of Ghana, but limited data at higher rainfall levels introduces uncertainty into these observations. In the Upper East and Ashanti regions, increased rainfall generally appears to have a smoothing effect that tends to increase malaria caseloads, though this trend becomes less certain at rainfall levels of 150 mm and above for the Upper East, and 300 mm and above for Ashanti (Fig 4 and S4 Fig 1D). Conversely, in the Northern and Central regions, higher rainfall levels correspond to a decrease in malaria caseloads, but the trend becomes increasingly uncertain at rainfall levels of 100 mm and above (S4 Fig 1B and 1H). Generally, these findings showcase the complexity of the relationship between malaria cases and rainfall, with regional variations and data limitations at higher rainfall levels contributing to the uncertainty in these trends.

The pattern of average temperature reveals contrasting trends, emphasising the complex and region-specific nature of the relationships between temperature and malaria cases. The effect of average temperature on malaria is more prominent in the Eastern and Volta regions compared to other regions, however the smoothing effect of average temperatures on malaria cases over the study period shows differing patterns between these two regions (Fig 4 and S4 Fig 1A–1I).

In the Eastern region (S4 Fig 1E), the smooth function plot reveals a consistent decrease in malaria caseloads as average temperature increases, with confidence intervals becoming narrower at 27–28 °C. In contrast, the Volta region (S4 Fig 1F) demonstrates a more nuanced relationship—as average temperature increases, the effect on malaria cases initially rises, following a quasi-linear trend until it reaches a peak at around 28 °C, after which the effect begins to decrease. This pattern suggests an optimal average temperature for malaria cases in the Volta region, which aligns with findings in other regions such as Ashanti, Central, and Western, though each region has its own distinct optimal temperature range. For instance, in the Ashanti region, the peak effect occurred at 27 °C, while the Central and Western regions exhibit their unique optimal average temperatures of around 26–27 °C for malaria cases.

The GAM results in Table 3 show significant temporal variations in malaria cases regarding the interaction effects of long-term trend and seasonality in all regions of Ghana, except for the Eastern region. This result can be interpreted to mean that the impact of seasonality on the dynamics of malaria cases in these regions is not static but changes over time [58], possibly due to factors such as climate change or evolving control measures. Furthermore, the interaction effect between the meteorological variables (rainfall and average temperature) shows varying significance across the various regions of Ghana. Notably, in the Eastern region, this interaction effect shown in Table 3 is significant (*p* < 0.05), indicating a strong combined influence of rainfall and temperature on malaria cases in this region.

The selected GAMs presented in Table 3 were assessed for model adequacy by comparing the fitted and observed data, examining the relationship between response and fitted values, and analysing deviance residuals for each regional model. These assessments indicated that the GAM assumptions were met (S5 Fig 1A–1C). Fitted values closely aligned with observed data, suggesting good model fit (S5 Fig 1C). The relationship between response and fitted values demonstrated appropriate patterns, further supporting model validity (S5 Fig 1B). Additionally, deviance residuals for each regional plot showed no concerning patterns or violations of model assumptions (S5 Fig 1A).

## Discussion

This study used temporal GAMs to assess the influence of weather conditions on malaria cases across the ten former regions of Ghana from 2012–2023. The analysis revealed complex temporal patterns influenced by seasonality, long-term trends, and weather conditions. Importantly, the seasonal patterns of malaria cases align closely with the peak case periods in various regions, though notable differences exist between regions.

In the regions constituting the Coastal and Transitional Forest zones, two peak malaria seasons were observed, consistent with previous studies [12, 55, 59–61]. These regions exhibit perennial and intense malaria caseloads, with slightly higher caseloads during the wet season. The malaria case patterns, however, show marked differences across the diverse ecological zones of Ghana. In contrast, the regions in the Guinea Savannah zone demonstrate a single peak season, coinciding with their long-wet season, followed by a pronounced decrease during the extended dry period. This more marked seasonality in the regions of the Guinea Savannah zone, relative to the Coastal and Transitional Forest zones, corroborates findings from a similar study in a comparable setting [32]. These distinct seasonal patterns across ecological zones demonstrate the spatial heterogeneity of malaria case dynamics in Ghana, emphasising the need for region-specific intervention strategies.

The interaction between long-term trends and seasonality was significant in most regions, except Eastern. In a similar study in Northwest Ethiopia, which evaluated seasonal and spatial variations of malaria transmission, space-time interaction effects were identified to have a higher contribution in explaining the temporal distribution of malaria transmission in the Amhara region [36]. This interaction effect observed in our study implies that the seasonal patterns of malaria cases are not static but change over time, illustrating the dynamic nature of malaria epidemiology. Such temporal complexity necessitates adaptive and responsive malaria control and elimination strategies that can anticipate and address changing seasonal case patterns.

This study established that the relationship between monthly malaria cases and meteorological factors varied across regions, confirming the complex interplay between climate and malaria cases in Ghana. The impact of rainfall on malaria cases demonstrated significant regional differences. The Upper East region showed statistically significant effects of rainfall on malaria caseloads. Other regions displayed less pronounced impacts. According to [62], rainfall can have both positive and negative effects on malaria transmission, depending on its temporal profile and variability. This complexity was evident in our findings, suggesting potential nonlinear interactions that differ by location.

Our analysis revealed increasing trends with some fluctuations in malaria caseloads for some seasons across various regions. However, these trends warrant cautious interpretation. An increasing trend in reported cases does not necessarily indicate a worsening malaria burden [31]. Several factors could contribute to this observed pattern, including improvements in the DHIMS2 data capture system [63], increased access to healthcare services [64], and changes in healthcare-seeking behaviour among the population. These factors could lead to better detection and reporting of malaria cases, rather than an actual increase in disease incidence. This points to the importance of considering both epidemiological trends and changes in health system capacity when interpreting long-term patterns in reported malaria case data.

The relationship between malaria cases and rainfall was found to be nonlinear and varied across regions. In Upper East and Ashanti, increased rainfall corresponded with a positive effect on malaria caseloads, albeit with increasing uncertainty at higher rainfall levels. This finding aligns with historical analyses reporting significant increases in malaria vector populations during the onset of the rainy season [65]. Conversely, the Central and Northern regions displayed a decrease in malaria caseloads with higher rainfall, though this trend also became less certain at higher rainfall levels. These findings suggest that the impact of rainfall on malaria cases is context-dependent, likely influenced by local factors such as drainage systems, mosquito breeding site management, and human behaviour. Furthermore, torrential rainfall may increase the flight range of mosquitoes when there is an increase in stagnant waters, potentially expanding their reach up to 1.7 *km* from breeding sites [66].

However, the observed regional variations in malaria cases may not be solely attributable to climatic conditions, as specific interventions and contextual factors may confound the expected relationship between meteorological variables and malaria cases. For instance, the comparatively low malaria cases observed in Greater Accra is likely influenced by urbanisation, which is associated with socio-economic and environmental changes that can reduce malaria cases, rather than being primarily driven by rainfall patterns [67]. Similarly, the low caseloads observed in the Upper East and Upper West regions may be partly attributable to IRS programmes implemented in these regions since 2011/2012 [68], which would have suppressed malaria cases independent of climatic conditions. Notably, the subsequent reduction of IRS coverage in Upper East to only three districts [69] may have contributed to the persistence of relatively low but not further declining caseloads in that region, as the partial withdrawal of a highly effective intervention would be expected to attenuate rather than eliminate its protective effect. These intervention-related and contextual factors represent important confounders that the GAM framework, while flexible in capturing nonlinear climate effects, was not designed to explicitly account for, and they should be carefully considered when interpreting regional differences in malaria caseloads.

Temperature significantly influences malaria cases, with distinct patterns observed in the Eastern and Volta regions. In the Eastern region, malaria cases decreased as temperatures rose, with most cases reported around 27–28 °C. Conversely, in the Volta region, malaria cases initially increased with temperature, reaching a peak at 28 °C, then declined. These findings suggest that optimal temperature ranges for malaria case burden vary by region. Also, the findings align with previous studies reporting peak transmission between 17 °C and 34 °C, typically around 25 °C [70, 71]. Regional variations indicate the critical role of local environmental factors and mosquito-parasite interactions in determining these optimal ranges. Understanding these dynamics is crucial for predicting and managing malaria outbreaks in response to temperature changes, underscoring the necessity for region-specific malaria control and prevention strategies.

As noted, previous studies have found malaria transmission to be driven by meteorological factors such as rainfall and temperature [54, 72–74]. Studies have also shown the effects of the interactions between these factors [75], however with some contradictory results [76–78]. Our study revealed that significant interactions between rainfall and temperature contributed to malaria cases, specifically in the Eastern region of Ghana. This finding suggests that, in this region, these factors together may create favourable conditions for mosquito breeding and malaria case burden. The results highlight the potential importance of considering the combined effects of multiple meteorological factors—rather than examining them in isolation—when assessing malaria risk and designing control measures for this region.

## Strengths and limitations

This study’s novelty lies in providing insights into regional malaria case patterns across Ghana. The comprehensive analysis of meteorological factors, particularly the nonlinear relationships between rainfall, temperature, and malaria, contributes valuable knowledge to the field of malaria epidemiology in Ghana. These findings can inform targeted interventions to reduce malaria cases by enabling health authorities to anticipate and prepare for high-risk periods based on regional weather patterns. For instance, regions could intensify preventive measures such as bed net distribution and IRS during periods when meteorological conditions are most conducive to malaria cases. Additionally, this region-specific understanding can help optimise the timing and allocation of resources for malaria control programmes, potentially improving their effectiveness and efficiency in reducing malaria cases across different areas of Ghana.

This study, however, has limitations. Firstly, it relies on confirmed malaria cases, which may be biased due to variations in access to healthcare and healthcare-seeking behaviour. Regarding data representativeness and completeness, all apparent missing entries were cross-checked with the respective data providers, and no missing data were identified at the point of analysis. Monthly records were subsequently inspected for implausible values and reporting inconsistencies, none of which were detected. While these checks indicate that the dataset was sufficiently complete and internally consistent for the purposes of this study, it is acknowledged that routine surveillance systems such as DHIMS2 are inherently subject to data quality challenges, including differential facility reporting completeness across regions and over time. Furthermore, variations in testing rates — reflecting differences in the availability of RDTs and microscopy services across regions and study periods — were not adjusted for in the analysis. Although these variations were not found to materially affect the overall analytical findings given the consistency of malaria patterns observed across regions, they may influence the comparability of confirmed malaria case counts and should be considered when interpreting regional differences. Secondly, socio-economic factors and vector surveillance data were not included in the study, which might limit the understanding of all regional potential determinants of malaria cases. Also, while covering a significant time frame (2012–2023), the study might not capture long-term climate change impacts on malaria patterns in the country. To address these limitations, our future studies will explore mathematical models that incorporate the issues raised, aiming to provide a more comprehensive understanding of malaria cases in Ghana.

## Policy implications

The findings of the study demonstrate the necessity for region-specific malaria elimination strategies in Ghana, tailored to the unique weather conditions and malaria case patterns of each region. Policymakers should prioritise developing adaptive and responsive interventions that account for the observed temporal variations in malaria cases, particularly the distinct peak seasons and nonlinear relationships with rainfall and temperature.

In regions like the Upper East and Ashanti, where increased rainfall is associated with higher malaria cases, enhanced vector control measures, such as improved drainage and mosquito breeding site management, should be implemented during rainy seasons. Conversely, in regions where higher temperatures or rainfall are associated with reduced malaria cases, efforts could focus on maintaining these conditions through environmental management and infrastructure development.

Additionally, the dynamic nature of malaria case patterns shown in this study calls for continuous monitoring and the integration of real-time climate data into malaria prediction models and early warning systems. Incorporating these elements would allow for more accurate and timely malaria outbreak predictions that account for region-specific climatic conditions, thus enabling more proactive regional interventions. Consequently, this approach can facilitate timely responses to emerging outbreaks and support more efficient allocation of resources. Lastly, collaboration between meteorological services, public health agencies, and local communities will be crucial in designing and implementing these tailored interventions, ensuring they are both effective and sustainable.

## Conclusion

The study revealed that combined effects of meteorological factors, rather than individual impacts, better explained malaria case patterns, particularly in the Eastern region. These findings emphasise the need for region-specific intervention strategies that account for local weather patterns and malaria cases. Our findings highlight that policy measures should focus on adaptive, region-specific interventions to effectively address the unique climate conditions driving malaria cases in each region. To this end, a key recommendation emerging from this study is the systematic integration of meteorological data into the National Malaria Data Repository (NMDR). Routinely linking climate variables with malaria surveillance data within the NMDR would enable health authorities to continuously monitor the impact of climatic conditions on malaria cases at the regional level, strengthen evidence-based decision-making, and support the timely deployment of targeted interventions during high-transmission periods. Future mathematical modelling studies incorporating regional variations and meteorological interactions, intervention coverage data, socio-economic factors, and vector surveillance data could further enhance our understanding of malaria cases in Ghana and inform targeted control measures.

## Data Availability

The malaria surveillance data (District Health Information Management System II [DHIMS2]) and meteorological datasets analysed in this study are available from the Ghana Health Service (National Malaria Elimination Program) and the Ghana Meteorological Agency (GMeT), respectively, upon reasonable request. Access to these datasets requires institutional permission due to national data governance policies. All code used in this study is openly available in the GitHub repository GAM?MAP (https://github.com/EdwardAkurugu/GAM-MAP). The repository includes scripts for data preparation, model specification, fitting, evaluation, and visualisation within the GAM framework. The interactive R Shiny dashboard developed for this study is accessible at https://masha-app.shinyapps.io/gam-map, providing a user?friendly interface for malaria analytics and visualisation.

https://gadm.org/download_country.html

https://ghs.gov.gh/

https://www.meteo.gov.gh/

## Ethical considerations

The ethical approval for the study was obtained from the Ghana Health Service Ethical Review Committee [**GHS-ERC: 019/04/24**] and the Human Research Ethics Committee of the University of Cape Town [**SCI/01798/2025**]. The access and permission to use the DHIMS2 malaria data and meteorological data were granted by Ghana’s National Malaria Elimination Program and the Ghana Meteorological Agency.

## Availability of the data and materials

The meteorological and malaria datasets used for this study can be obtained from the GMet and NMEP of the Ghana Health Service, respectively, upon request.

## Code availability

All code used in this study is openly available in the GitHub repository **GAM-MAP** at https://github.com/EdwardAkurugu/GAM-MAP. The repository contains scripts for data preparation, model specification, fitting, evaluation, and visualisation within the GAM framework. The code is released under the license specified in both the repository and the manuscript, and it can be freely accessed to reproduce or extend the analyses presented here.

## Software availability

The interactive dashboard developed for this study was implemented in **R Shiny** and is openly accessible at https://masha-app.shinyapps.io/gam-map. The GAM-MAP dashboard provides a user-friendly interface for malaria analytics and visualisation. Users can explore the model outputs, interact with the data, and reproduce the analyses through the application under the terms specified on the site.

## Author contributions

**Edward Akurugu:** Conceptualisation, methodology, investigation, software, formal analysis, writing – original draft, data curation, and visualisation.

**Timothy Awine** and **Baba Seidu:** Data curation, supervision, writing – review.

**Sheetal Prakash Silal:** Supervision, funding acquisition, resources, writing – review.

**Paul Hilarius Asiwome Kosi Abiwu, Nana Yaw Peprah, Wahjib Mohammed, and Paul Boateng**: Data curation, validation, writing – review.

## Acknowledgement

We acknowledge the Modelling and Simulation Hub, Africa (MASHA) at the University of Cape Town for providing funding through the Malaria Modelling and Analytics: Leader in Africa (MMALA) project to support the PhD studies of the first author (Edward Akurugu). We thank collaborators such as the Clinton Health Access Initiative (CHAI) in South Africa and the Southern African Development Community Elimination 8 (SADC E8) for their technical support during data acquisition. The opinions, findings, and conclusions expressed in this study are solely those of the authors and do not necessarily reflect the views of MASHA or its collaborators. We further acknowledge Dr. Keziah Laurencia Malm for her guidance and assistance during the acquisition and collation of data from the NMEP.

## Supporting information

**S1 Text. GAM description and equations.**

**S1 Fig. Conceptual flowchart of the regional GAM building process algorithm**

**S2 Fig. Patterns of monthly regional aggregated malaria cases for 2012 to 2023.**

**S3 Fig. Malaria cases series decomposition (trend, seasonal, and remainder) by LOESS for each region from 2012 to 2023.**

**S4 Fig. Regional partial effects of covariates (long-term trend [time], seasonality [months], rainfall, average temperature, interactions between long-term trend [time] and seasonality [months], and interactions between rainfall and average temperature) on malaria cases, 2012 to 2023.**

**S5 Fig. Regional selected GAMs diagnostics.**

